# “A baby is like an empty USB. Whatever you do, they store it”: a qualitative study of caregiving practices and beliefs in Soweto, South Africa

**DOI:** 10.1101/2024.11.06.24316849

**Authors:** Tom Palmer, Nokuthula Nkosi, Molebogeng Motlhatlhedi, Audrey Prost, Jolene Skordis, Lu Gram, Shane A Norris, Neha Batura, Catherine E Draper

## Abstract

The importance of nurturing care for children’s development has been emphasised in the global literature. Although societal risk factors which may hinder nurturing care in South Africa have been extensively documented, the goals, beliefs and practices of caregivers in infancy have received less attention in the literature. This qualitative study was conducted with mothers of children aged 6 to 10 months in Soweto. Data were collected during focus group discussions held in May 2023 with 26 mothers, then analysed using thematic analysis. We found that mothers view themselves as playing an active role in their children’s learning and development, and are broadly aware of the importance of nurturing care. Our findings also emphasise that nurturing care is bidirectional, and based on the relationship between caregivers and children, both of whom should be viewed as individuals. This study contributes to relatively limited literature on infancy in South Africa and can help to inform context-appropriate interventions to improve early childhood development in comparable settings.

## Introduction

The statistic that around 250 million children under five years of age in low- and middle-income countries are at risk of poor development [1], based on proxy indicators of stunting and poverty, has been extensively cited in the early childhood development (ECD) literature. The Nurturing Care Framework (NCF) was launched at the 2018 World Health Assembly and outlines a roadmap for how those children can be targeted by interventions [2]. Within the NCF, nurturing care is identified as the main way to improve ECD, and caregivers (typically parents but potentially also grandparents and wider extended family) are therefore viewed as central to improved ECD. In South Africa, modelled estimates suggest that only 20.5% of children aged 3-4 years receive minimally adequate nurturing care [3]. Societal risks to effective caregiving and child development, including urbanisation, poverty, violence and single-adult households have been extensively documented [4–12].

Within this context, the beliefs and practices of South African caregivers during infancy have received less attention in the literature. Historical literature on infancy in South Africa has tended to focus on protection and survival. De Wet [13] gives an overview of classical ethnographies describing traditional protective practices and shows how contemporary use of medicines in infancy in Soweto, a low-income urban area in Johannesburg, could be seen as adaptations of various folk healing traditions. Richter [14] suggested that all caregivers across cultures engage in protective practices and rituals during infancy that are related to their their environment, while highlighting the lack of research on infant care in South Africa.

More recently, studies in Khayelitsha, a low-income urban community in Cape Town, found low levels of sensitivity during play interactions and high levels of intrusive parenting, where caregivers impose their own agenda and heavily interfere with infant behaviours [15,16]. However, five recent qualitative studies in urban South Africa have explored caregiver beliefs about ECD, and all found that caregivers were aware of the importance of what could broadly be described as nurturing care [17–21]. Most of these qualitative studies focused largely on barriers and enablers to effective caregiving, while only one focused specifically on practices in infancy [21].

A key strategy for targeting caregivers is parenting interventions which aim to change their behaviour or beliefs, typically through home visits or group interventions [22–29]. However, concerns have been raised regarding the assumed developmental universalism implicit in the rationale for parenting interventions, as well as the risk of promoting goals for childrearing that are inconsistent with local parental goals, and which may have unintended negative consequences for children, families and communities [30–32]. Additionally, such interventions often focus entirely on the responsibility of mothers for caregiving, neglecting diversity in caregiving arrangements and practices globally [33].

Parental ethnotheories are the cultural belief systems of parenting and child development held by caregivers [34]. These beliefs are important influences on caregiving practices [34] and caregivers’ psychology [35]. Understanding parental ethnotheories is crucial for designing interventions to improve ECD [35]. Insights into caregiver beliefs about developmental goals and the practices used to achieve them is essential for parenting interventions to successfully promote behaviour change. In this study, we explored the caregiving goals, practices and beliefs amongst mothers of infants aged 6-10 months in Soweto, South Africa, to gain an in-depth understanding of ECD in this setting. This study contributes to relatively limited literature on infancy in South Africa and can help to inform context-appropriate interventions.

## Methods

### Context

The *Bukhali* trial is being conducted in Soweto, a predominantly low-income urban area of Johannesburg, South Africa, amongst women aged 18-28 years. The trial tests a complex continuum of care intervention beginning in preconception and continuing throughout early childhood. The intervention and trial design are described in detail in the trial protocol [36]. The present study was conducted with mothers participating in the *Bukhali* trial who had a child aged between 6 and 10 months. This age group was selected given relatively limited research on infancy in South Africa, and as this is a key, mostly pre-verbal, developmental stage, where awareness on early learning could be lower than for older children, with research in some settings suggesting nurturing care is more limited in the first year of life [37]. Our study aligned with the trial process evaluation objective of understanding context and lived experience of participants [38].

### Ethics statement

Ethical approval was received from the Human Research Ethics Committee (Medical) at the University of the Witwatersrand (M190449) in South Africa and from the University College London Research Ethics Committee (14657/003) in the United Kingdom. Prior to data collection, all participants provided written and verbal informed consent to participate in FGDs, and for the FGDs to be recorded.

### Sampling and data collection

Participant recruitment and data collection took place from 1^st^-5^th^ May 2023 through focus group discussions (FGDs), which were facilitated by a researcher with an undergraduate degree and extensive experience collecting qualitative data in this setting (MM), in addition to one local (NN) and one non-South African (TP) note-taker. Both local researchers were conversant in local languages. In total, 26 mothers participated in the study. A total of four FGDs were conducted ranging from 80 to 114 minutes in duration, each with between five and eight participants. Mothers ranged from 20 to 29 years of age, and their infants ranged from 6 to 10 months of age. For most mothers (n=17) this was their first child, while eight mothers had one older child, and one mother had two older children. Only four mothers were employed at the time of data collection. The most common home language amongst participants was isiZulu (n=14), followed by Sesotho (n=7), Setswana (n=2), Venda (n=1), Tsonga (n=1) and Xhosa (n=1).

FGDs were conducted at a research centre within Chris Hani Baragwanath Hospital in Soweto. A ZAR150 transport reimbursement and refreshments were provided for participants. All discussions were audio recorded. Verbatim transcription was conducted, followed by translation from local languages to English. Topic guides and written guidance for facilitators were developed prior to data collection, with an initial draft developed by TP and refined through contributions from CD, LM, NN, AP and JS. Questions explored caregiver goals, aspirations, behaviour, and beliefs around contributing factors to childhood development. Although the topic guide covered topics broadly related to the NCF, specific questions were not directly linked to elements of the NCF. Questions were generally broad and open-ended, though specific questions were asked related to talk, play and school readiness. The topic guide is provided in an appendix.

### Data analysis

Data were analysed by TP using a combination of inductive and deductive qualitative thematic analysis [39,40], moving between the inductive insights from the data and the deductive framework, or theory, as outlined in the NCF. This approach was chosen as a “pure” inductive approach was not deemed possible, as both analysis and the topic guide were inevitably influenced by existing literature and theory, particularly given that this research took place within the context of the latter stages of a PhD on the topic.

Analysis took place in two stages. First, an initial codebook was developed deductively based on the NCF, reflecting the influence this document may have had on the topic guide (and the wider ECD literature). This included the five components of nurturing care (security and safety, responsive caregiving, opportunities for early learning, good health, adequate nutrition) five components of development (physical, social, emotional, cognitive, language) and four components of an enabling environment (supportive services, enabling policies, empowered communities, caregivers’ capabilities) outlined in the document [2]. Social and emotional development were combined in the deductive codebook, given substantial overlap in these two developmental domains.

After data familiarisation, transcripts were indexed to the broad codes from the deductive codebook [41]. This anchored the content to the NCF, by seeking data related to each element of the NCF, reflecting that the topic guide structure may have been influenced by the NCF. Detailed research notes were written during this process, including beginning a list of preliminary analytic codes as insights occurred.

During the second stage of analysis, more refined analytic (i.e. inductive) subcodes were then applied to indexed sections of the transcripts within each of the broad deductive domains. These analytic codes therefore combined both emergent findings and existing theory. Analytic codes were further refined through multiple rounds of reading through index codes. Codes were then synthesised into themes which captured meaningful patterns in the data.

### Reflexivity

This research forms part of TP’s PhD, which focuses on the beliefs and practices of caregivers of young children, including prior research in South Africa and India. TP is a British male, and played a role in the construction of qualitative data by leading topic guide development and data analysis. TP has a background in economics, and had prior experience in qualitative data collection and analysis, including in South Africa. South African co-authors NN, MM and CD have extensive experience of ECD research in low-income settings throughout South Africa, and provided substantial contributions to topic guide development, data collection, notetaking, interpretation of findings, and finalising the manuscript.

## Results

Three themes are reported from our analyses: (1) Mothers’ goals and roles; (2) Nurturing connections; and (3) External influences on caregiving practices. These themes capture mothers’ diverse experiences of caregiving in this setting. Each theme is illustrated with relevant extracts from the data.

### Theme 1: Mothers’ goals and roles

Overall, mothers acknowledged the active role they played in their child’s learning and development, and were broadly aware of the importance of nurturing care. Mothers described various goals and aspirations for their children which influenced their caregiving practices. Separate subthemes are presented below for infancy and early childhood, reflecting changes in roles and goals over the first few years of the child’s life.

> *“A baby is like an empty USB. Whatever you do, they store it.”*
>
> *“It’s folded while it’s still wet mama. You train them now*.*”*
>
> *“Every day you are teaching your child something new*.*”*

#### Infancy: An active child with a good appetite

Gaining weight and physical growth were the most frequently mentioned indicators of current healthy development (i.e. during infancy), and mothers described feeling happy when their child’s weight increased. Children eating well was therefore a key goal for mothers at this age, and mothers described feeding as being a potential source of stress. Another common goal expressed by mothers was for their child to be “*active*”. This appeared to encompass a combination of physical, socioemotional and cognitive developmental domains, and was considered an overall indicator of a child’s healthy development. Mothers reported playing with their children, including with toys, though it was also very common in this setting for infants to play with older children from the household or neighbourhood. Some mothers explicitly noted developmental reasons for play, while others described play as something that mothers and children enjoyed doing together, linked to a goal expressed by some participants of making their child happy.

> “*For me good child development is when the child is gaining weight. So I like children who are chubby and stuff. So every time I put my baby on the scale, when it goes up I get excited. And every time his weight goes up it shows that the baby is eating well. He’s alright, there’s nothing wrong with him.”*
>
> *“I need my child to have an appetite and to eat and be active. Yes, I like an active child*.*”*
>
> *“I want them to be active, to be a child that likes to explore, when they see something on the floor they are interested and they want to see what it is and what it does*.*”*
>
> *“Like, I believe that actions speak louder than words, and seeing my baby happy gives me enough proof that, damn I’m doing a good job despite whatever I’m facing. I’m doing a good job and nobody can take that away from me*.*”*

#### Early childhood: A normal child

Looking ahead beyond infancy, mothers commonly expressed future goals for their child’s socioemotional development, including liking, respecting and accepting other people. This could broadly be summarised as hoping for their child’s successful social adjustment. Some mothers also described their hopes for their child to be “*norma*l” or “*like other children*”, highlighting the value placed on fitting in. Very few participants mentioned aspiring for their child to be intelligent. Mothers were more likely to mention aspirations for qualities such as calmness, self-knowledge and independence. These qualities formed a broader view of self-mastery, rather than narrowly focusing on cognitive development. However, mothers were also asked directly to reflect on what helps make children ready for school, and acknowledged their role in preparing children for school, with responses largely focusing on various forms of knowledge that children should be taught, such as basic numeracy and literacy, or naming colours and shapes.

> *“I want my child to like people. I want them to accept people, who they are and their situations. If they visit someone and they find that at that place they are eating cabbage, they should eat cabbage with them, they should not be judgemental*.*”*
>
> *“I want my child to be in tune with themselves. I feel like if you’re not in tune with yourself, to be honest, nothing is gonna go okay*.*”*
>
> *“One thing that we tend to forget is that you as the parent you are the child’s first teacher. Not at the school where they are qualified to teach the child. You as the parent you are your child’s first teacher, because, if you notice, we teach our kids everything they know. They just grow to add on what we taught them already*.”

### Theme 2: Nurturing connections

This theme captures that nurturing care can be seen as an outcome of relationships between caregivers and children, rather than simply something caregivers provide unidirectionally to their children. That is, while mothers influence child development, their own lives are in turn influenced by children through a bidirectional relationship. Mothers reported enjoying interacting with their child and seeing their development, and the importance of a strong connection or bond with their child was frequently discussed. Mothers spoke about these relationships positively and described getting along with their children and loving each other. Mothers’ descriptions of their daily interactions and routines were frequently in line with what could be described as responsive care. Mothers emphasised that all babies were different and described understanding their babies and their needs and responding accordingly. The importance of caregivers being “*calm*”, “*patient*”, “*friendly*”, and giving love to their babies was frequently expressed. Despite the inevitable challenges of parenting, and many mothers describing feelings of “*stress*” and insecurities, these positive relationships appeared to positively impact nurturing care, and enrich mothers’ lives.

*“But it’s such a nice experience to see someone, your little one, growing. Their first noises. It’s nice. Enjoy every moment*.*”*

*“Being a mother is a really wonderful thing, seeing your child’s development as they get older, that bond. You should focus on that a lot, more than the circumstances that you’ll be faced with.”*

The routines described by mothers were also responsive to their children’s individual needs, including preferences for food and sleep. Being responsible for providing nurturing care of course affected mothers’ lives, most notably including a lack of spare time and sleep. Most frequently, mothers described difficulties in finding time for domestic tasks, while a few also mentioned that they lacked time to spend on themselves. For many, the only time available for domestic tasks was when their child was asleep, engaged with a screen, or in a walking ring. Walking rings were very widely used amongst participants, and most mothers reporting having one.

> *“You won’t be able to do washing while your baby is awake. You have to do it while they’re asleep. So that’s when I do washing properly at night. I wash, rinse, dry. Tomorrow morning wake up and I hang. Same thing like ironing. So now she’s messing up my sleeping patterns*.*”*
>
> *“For me, I feel like I don’t focus on myself like I focus on my child. I would wish to have time to go out with the girls and get some fresh air, you know? Like… cause like I once did, I went but I missed my child, so like I want to do things without him, let me just say that, like doing things without him there*.*”*

Mothers saw their pre-verbal children as interactive partners and acknowledged the role this had in shaping language development, including implicitly through negative opinions of “*baby talk*”. Mothers reported talking and interacting with their children from a very early age, sometimes starting in pregnancy. Some mothers also described how interacting with their children had benefits for themselves, such as feeling understood by their child. Again, this highlights the bidirectional nature of responsive caregiving and the importance of the caregiver-child relationship for nurturing care.

> *“I do talk to her. I speak proper because what I’ve realised, when you speak proper, it’s easier for the child to learn to speak quicker. Because sometimes baby talk… I’m not saying I’m against it, but then sometimes it delays the speaking of the child. So when you speak proper the child can learn quicker*.*”*
>
> *“Coz it was just the both of us. So I can’t just keep quiet. I just talk to him. ‘Hey nana how are you? Are you okay? What’s this? Okay. Let’s go. This, this*.*’”*
>
> *“As a first-time mom there’s a friend of mine. I told her, yoh, I’m going through a lot. Then she said “[Name]. Please stop telling me your problems. Tell your child your problems. Your child is your therapist*.*” And I was like “heh!!”. Fine I spoke to him, but I didn’t wanna talk to him about, “ey I’ve got a certain pain somewhere”, “someone is treating me badly*.*” And then I started doing that. And I actually felt better. Because now as much as he’s not responding, but the facial expressions and the gestures, they mean the world to me. It’s like I’m talking to God himself*.*”*

### Theme 3: External influences on caregiving practices

This theme reflects the varied external influences on caregiver behaviour, at family, community and societal level. Mothers reported various influences, both material and informational, that influenced their parenting and their child’s life and development. In terms of family structure, many participants lived in single parent households, with fathers often absent. Some participants mentioned that the father played a role financially but spent little time with their child. When this was the case, other family members, typically the maternal grandmother, often played a major role in supporting the mother in raising the child. However, some participants also described supportive fathers who played a substantive role financially, emotionally, and physically.

> *“It is painful that the father of the child isn’t close to the child anymore*.*”*
>
> *“The fathers of the babies leave us, and then you take that anger and point it towards the child, then the child does not grow up well. What I can say is if you have your own child is to be focused on the child and not focus on all the other stuff that’s happening around*.*”*
>
> *“The father of my child is present. Sometimes I even wish that he can go away for a day*.*”*

Mothers reported receiving advice from elders and others in the community on caregiving practices and traditional beliefs. This was generally appreciated and seen as a useful source of information on parenting. Some intergenerational differences were acknowledged by participants. For example, one participant reflected on the ubiquity of branded baby foods, and how this differed from the previous generation. In general, branded baby foods were frequently referred to where food and nutrition were discussed, as were walking rings during discussions of daily routines, suggesting commercial influences on caregiver behaviour.

> *“They know better…Some way, somehow, yeah they know better. I do listen to elder people. Even though my child came before his time. But I listen to them*.*”*
>
> *“So older people are more experienced than us. Because with us we take the easy way out. We give the children solid foods that just need you to mix water in them. But with them, they’re the type to cook pap and beans. So I think their advice does help us, even though sometimes we can make that Purity or that Nestum*, it’s the easy way out. With them they had to cook pap and stews. And then they’ll say okay, at a certain age give them these kinds of food and they’ll be alright. And then along the way when you give them this it treats them well. So along the way their advice helps us”*
>
> *\*Widely available baby food brands in South Africa*

Some caregivers appreciated that their community showed an interest and played with their child, such as when they were out shopping. However, others remained wary of strangers, and even family members. A few mothers reflected on the dangerous environments in which they live, and the possible risks this might have for child development. This also included a few allusions to a lack of trust in strangers due to the potential harms of witchcraft, which might cause children to fall ill.

> *“My environment is very bad… you don’t know who actually loves you and, you know, where the danger is with your child… So, the environment I would say it’s not the best for the child’s development, you understand? Because I, for one, I don’t trust anyone with my child. Anyone. Even my mother-in-law*.*”*
>
> *“I believe as people we believe in different things. Some of us are Christian. Some of us believe in religious things like tradition. So people toughen up to protect themselves with whatever they believe in. So you can only imagine if they’re going to hold your child and their energy is weighing them down and when they sleep it’s a problem. You just don’t allow anyone to hold your baby. It’s one of those things*.*”*
>
> *“We are people of colour, you know these things. Someone will tell you “grandmother so and so is doing this”. So you obviously wouldn’t allow your baby to be exposed to such, you understand?”*

Most mothers reported using screens, and cartoons such as *Cocomelon* were widely watched. Most frequently this was to keep their child entertained or so that they could complete domestic tasks, but some mothers also mentioned educational benefits of television. Only one mother mentioned reading books with their child, and described this in a way that suggests it is not common in this context. Only a couple of participants raised explicit concerns about the benefits of children using phones, or the extent of their phone use. This included one mother who viewed phones as distracting her from playing with her child. However, overall, screen time has a significant influence on the daily experiences of children in this setting.

> *“We spend a lot of screen time, like we have a lot of screen time… Okay, I tried reading him a book, my mom even said “you’re doing white people things”, and my son was just sitting there looking at me like… I don’t know if he understood, but he looked confused”*
>
> *“When my child cries, the moment he cries, like maybe he’s sitting down, I’ll take the phone and open it and he is dead silent and he just smiles. So I try to avoid it most times because he will become dependent on it”*
>
> *“Sometimes I am lazy to play with my child…Sometimes on social media there would be interesting stuff being posted. You will find that on Facebook or TikTok things are happening*.*”*

Policies or services were not frequently referred to by caregivers, perhaps reflecting the fact that families are seen as responsible for child development, or a broader lack of awareness of their entitlements and the wider impact of policy on the environment in which they are raising their children. Only one participant referred to receiving a grant, though questions were not asked specifically on finances. The only services that mothers mentioned were clinics and creches or preschool. Thus, policies and services do not appear to be major external influences on caregiver behaviour in this setting. Clinics were mentioned by a few participants as providing support by monitoring their child’s health or physical development, though it is worth noting that the FGDs took place within this setting. Creches were mentioned by several participants as places that taught children and enabled mothers to work, while some participants also mentioned the benefits of socialising with other children. Mothers had differing views over the quality of available creches, and the level of attention that children would receive, with some viewing creches as just a necessity, whereas others perceived creches to have educational benefits. However, creches were not a substantial influence on caregiver behaviour in this study, given that most mothers were not currently employed, and had young infants.

> *“Have you ever seen a child who has stayed at home for a long time? The moment they go to school, and they haven’t been to crèche before, they are very slow at school*.*”*
>
> *“For most crèches they’re just babysitting but they don’t teach them*.”
>
> *“Some crèches you cannot be sure that they’ll do the right thing. Accredited or not*.*”*

## Discussion

This article aimed to outline the caregiving goals, practices and beliefs of mothers of infants in Soweto, South Africa. We found that mothers view themselves as playing an active role in their children’s learning and development, and are broadly aware of the importance of nurturing care, in line with findings from other recent qualitative studies in urban South Africa [17–21]. We expand upon this literature by providing further insight into the goals and practices of mothers specifically during infancy, and reflecting on different developmental domains. Additionally, we considered the impact caregiving has on mothers.

Keller [42] outlines two “prototypical” parenting styles: proximal and distal. Proximal parenting is typically associated with rural subsistence settings and prioritises engagement through physical touch, while distal parenting is seen as characteristic of industrial or postindustrial middle class families, and is characterised by object stimulation, face-to-face exchanges, and vocalisations [43]. A study using observations of caregiving in Khayelitsha, Cape Town, concluded that infants in low-income urban communities in South Africa do not experience either of these prototypical models and noted a lack of understanding of childcare in such settings [44]. However, mothers in our study reported behaviours indicative of distal caregiving, including use of walking rings, verbal interaction, and goals for self-knowledge.

These findings could indicate shifting parenting styles in Soweto and other comparable settings. Another study in Gauteng province, South Africa, highlighted the impact of acculturation on caregiving practices, and also noted that face-to-face interactions were common in infancy [21]. An exploratory study in Alexandra, Johannesburg, found differences in maternal sensitivity scores depending on the choice of measurement scale, and highlighted the need develop indicators for context-specific manifestations of sensitivity [45]. We also highlighted that caregiving is not static through exploring the external influences on caregiver beliefs and practices. We found high levels of screen time amongst participants, while wider evidence from South Africa also suggests that average screen time for children is likely to exceed the levels recommended in the guidelines [46–48]. However, existing research in South Africa has primarily focused on screen time in the context of sedentary behaviour amongst older children, rather than as a risk factor during infancy. High levels of screen time, particularly when passive, may displace opportunities for interactive and responsive caregiving, thereby constraining child development [46,49–53].

As might be expected, mothers in our study frequently reflected on their lack of time and sleep, near universal experiences for parents that are likely to influence maternal mental health [54,55]. However, our second theme also emphasises more broadly that nurturing care is bidirectional, and based on the relationship between caregivers and children, both of whom should be viewed as individuals. Most theories of caregiver-child relationships emphasise how children and their caregiving environments have bidirectional, interdependent influences on one another [56,57].

These findings have implications for the NCF and the design of parenting interventions. In the NCF, improvements in developmental domains are framed in terms of the benefit to the child, as well as to wider society through supposed economic benefits. The perceived benefits to caregivers are not considered, though surely these developmental outcomes can be considered as the goals which guide caregiver behaviour, including provision of nurturing care. In terms of the benefits of positive relationships with their children, the NCF is limited to a single brief acknowledgment of the possibility that “*mutually enjoyable interactions create an emotional bond*”. The overall impression is left of nurturing care as purely outcome-oriented. Behaviour change initiatives may be more likely to succeed if they emphasise immediate affective outcomes, such as enjoyment or joy, rather than long-term rational outcomes, such as child development [58]. Mothers in our sample reported enjoying their relationships with their children, and it seems sensible that interventions emphasise this.

Our study is one of few known qualitative studies to explore caregiver beliefs, goals and practices during infancy in South Africa. However, this study has several important limitations. First, due to a need to comply with trial unblinding procedures, it was not possible to make comparisons between participants in the intervention and control arms of the *Bukhali* trial. Although the aim of this study was not primarily to assess caregiver knowledge, this could be the subject of future work using this dataset. Additionally, all participants were actively engaged with health services and therefore may not be generalisable to mothers in this setting. Finally, standard limitations of FGDs, such as potential adherence to socially acceptable opinions, apply. Ethnographic research may provide more insight into what people do rather than what they say, but is comparatively resource intensive.

## Data Availability

The authors do not have permission to share the data for this study, due to ethical concerns from the University of the Witwatersrand Human Resource Ethics Committee about sharing qualitative interview data outside of the research team. This is because the focus group discussions and the de-identified transcripts thereof contain potentially identifying and sensitive participant information. Data can be made available to interested researchers upon request to the HREC (Medical) at the University of the Witwatersrand.

## Notes

### Competing Interest Statement

The authors have declared no competing interest.

### Funding Statement

This work was funded through internal seed funding aiming to increase collaboration between University College London and the University of the Witwatersrand (Wits-UCL Research/Teaching collaborative activity Seed Fund 2022/23). The funder played no role in the design, analysis, or writing for this study.

